# Associations Between Physical Activity Intensity and Experience, Self-Regulation, and Self-Reported Interoceptive Accuracy and Attention

**DOI:** 10.1101/2025.05.06.25326015

**Authors:** Jesper Mulder, Jessica C. Kiefte-de Jong, Juriena D. de Vries, Marije T. Elferink-Gemser

**Author notes:** Contact information corresponding author: Jesper Mulder, Health Campus the Hague, Department of Public Health and Primary Care, Leiden University Medical Center, The Hague, The Netherlands, Turfmarkt 99, 2511 DP The Hague, The Netherlands.

## Abstract

Interoception may play a key role in healthy lifestyles due to activation of similar brain regions during physical activity (PA) and interoceptive tasks. Previous research concentrated on acute PA and single interoceptive domains (e.g., accuracy or attention). We examined how 1) varying PA intensities, 2) years of PA experience, and 3) self-regulation associate with self-reported interoceptive accuracy and attention.

The Interoceptive Accuracy Scale and Interoceptive Attention Scale were used in 1178 participants (mean age 52.13 ± 16.78, 52% female). The leisure-time activities domain of the International Physical Activity Questionnaire with additional in-depth questions was used to determine PA intensities and experience. Self-regulation was assessed using the Short Self-Regulation Questionnaire.

Log-normalized regression results indicated walking hours per week (M = 3.75 ± 4.17, β = 0.009, p = 0.028) and self-regulation scores (M = 109.36 ± 13.50, β = 0.262, p < 0.001) were significant determinants of self-reported interoceptive accuracy, while years of PA experience (M = 13.39 ± 15.39, β = −0.022, p = 0.005) and self-regulation (M = 109.36 ± 13.50, β = −0.147, p = 0.036) were significant determinants of self-reported interoceptive attention.

Walking hours associated with an increase in interoceptive accuracy, while more years of PA experience decreased interoceptive attention. Better self-regulatory skills increased interoceptive accuracy, and decreased interoceptive attention. Being able to filter relevant interoceptive information may be extremely important in health behavior. We encourage future studies to investigate whether individuals with better self-regulation are therefore more physically active, resulting in increased interoceptive accuracy.

**Highlights:**

- Adaptation to low intensity physical activity reduces sensitivity of bodily signals
- Years of experience in physical activity reduces interoceptive attention
- During vigorous intensity physical activity internal noise clouds interoception
- Better self-regulation and interoceptive accuracy increases bodily trust
- Filtering relevant interoceptive information may be important in health behavior

## 1. Introduction

Interoception, the perception of internal bodily signals, may play a key role in promoting a healthy lifestyle, specifically in engaging in physical activity (Mulder et al., 2025). During interoceptive accuracy tasks (e.g., heartbeat tracking tasks), brain activation is not only found in the insular cortex, anterior cingulate cortex, and prefrontal cortex, but also in the primary motor cortex and supplementary motor area (Critchley et al., 2004; Zarza Rebollo et al., 2019). The activation of these brain areas suggests a relation between motor regions in the brain and interoceptive pathways. When interoceptive signals are insufficiently or excessively perceived, this may lead to adverse health behavior. For example, when there is a mismatch between perceived fatigue and the actual physiological state during physical activity, this can result in hypoactivity (i.e., reduced participation in physical activity, which may lead to problems related to sedentary behavior) or hyperactivity (i.e., continuation of physical activity to the point of reaching dangerous physiological boundaries; Wallman-Jones et al., 2021). These mismatches could damage the body, therefore the brain regulates physiological needs before homeostasis is threatened (McMorris et al., 2018). There are indications of a bidirectional relation between physical activity and interoception: physical activity helps to tune internal models (i.e., the brain’s ability to predict and control movements based on prior experiences and sensory feedback), and better tuned internal models allow for better management of exertion (Seabury et al., 2023). This highlights the importance of the relation between physical activity and interoception, and understanding this relation can provide insights into how physical activity is initiated, regulated, and maintained.

Despite the suggested relation between motor and interoceptive brain areas, current research on the relation between physical activity and interoception is limited to single interoceptive domains or modalities, such as interoceptive accuracy and heartrate based tests (Abrams et al., 2018; Herbert et al., 2007; Kosa et al., 2021; Koteles et al., 2020; Machado et al., 2019; Teng et al., 2018), and/or acute physical activity protocols (Wallman-Jones et al., 2023; Wallman-Jones et al., 2022). Research distinguishes two domains: interoceptive accuracy, the ability to adequately interpret interoceptive signals, and interoceptive attention, the extent to which one pays attention to interoceptive signals. Different modalities can, for example, include the perception of respiratory sensations or digestive processes. A recent study argued that different interoceptive domains and modalities relate in different ways to several disorders, showing the need for research that includes both domains and several essential modalities (Schoeller et al., 2025). Moreover, to the best of our knowledge only one study has investigated the longitudinal effects of physical activity on interoception (Amaya et al., 2021). It was found that after two to three months of physical activity at moderate intensity interoceptive accuracy was found to increase, indicating the positive influence of repeated exposure to physical activity on developing interoceptive accuracy. Additionally, an individual’s physical activity background and experience may influence interoceptive responses: sporting background predicts interoceptive accuracy in rest and following physical activity, with students not majoring in sports courses showed increased interoceptive accuracy after physical activity compared to students majoring in sports (Wallman-Jones et al., 2022). These findings underscore the need for more comprehensive research on how different physical activity intensities, backgrounds, and levels of experience associate with various domains of interoception.

Self-regulation plays a key role in initiating, sustaining, and adapting to (regular) physical activity (van Genugten et al., 2017). Self-regulation commonly refers to cognitive and behavioral strategies that facilitate behavior adaptation, and comprises of meta-cognitive functions such as the ability to reflect, plan, monitor, and evaluate a goal-directed process as well as aspects of motivation and self-efficacy (Menting et al., 2019). Self-regulation of physical efforts is relevant not only for managing fatigue in people with long-term conditions (e.g. chronic pain conditions, cardiovascular diseases, neurological conditions, arthritis, cancer, and gastrointestinal conditions) and promoting sustained physical activity, but also for improving health-related quality of life (Barakou et al., 2023). Those with better self-regulatory behavior may be better able to perceive and interpret bodily signals, as they can identify what requires immediate regulation. Conversely, those with lower interoceptive accuracy may struggle with self-regulation, as they lack awareness of the most urgent physiological needs (Füstös et al., 2013; Georgiou et al., 2015; Herbert et al., 2007; Marshall et al., 2018). Thus, it could be that individuals with better self-regulatory skills also display better self-reported interoceptive accuracy, supporting engagement in sufficient physical activity.

The purpose of this study is to examine how varying intensities of physical activity (e.g., walking, moderate intensity, and vigorous intensity), physical activity experience, and self-regulation are associated with both self-reported interoceptive accuracy and attention. Our hypotheses are that 1) participation in walking, moderate intensity, and vigorous intensity activities incur positive increases in interoceptive accuracy and attention; 2) more years of experience in physical activity are positively associated with self-reported interoceptive accuracy and attention scores; and 3) self-regulation scores are positively related to self-reported interoceptive accuracy.

## 2. Methods

### 2.1. Population

Participants in the current study were from the general Dutch population, recruited through a Dutch internet panel administered by research agency Flycatcher (www.flycatcher.eu). Through Flycatcher’s ‘double-active-opt-in’ participants voluntarily and actively indicated their willingness to participate in this study. Flycatcher’s panel consists of 20.000 members from the Dutch general population, giving a fair representation based on sex, age, region, and socio-economic status. Flycatcher meets high quality standards and is ISO-certified, and offers participants points to be spend on (charity) gift vouchers for completing surveys. Participants had to meet the following criteria to participate in the study: 1) the subject was at least 18 years old, and 2) the subject had a clear understanding of the Dutch language. Sample size calculations for linear multiple regression analyses using G*Power 3.1 (Faul et al., 2009) suggested a minimum sample size of 1240 participants, based on an effect size of 0.02, alpha of 0.005 (0.05 divided by 10 predictors in the original study protocol), and a power of 0.8. Participants were contacted via email to take part in the study.

### 2.2. Measures

#### 2.2.1. Self-reported interoceptive accuracy

The Interoceptive Accuracy Scale (IAS; Murphy et al., 2020) is a questionnaire constructed to include items relating to physical sensations that have either been described as interoceptive or are associated with activation in the insula, an area commonly associated with the processing of interoceptive signals. An example of a question in this questionnaire is “I can always accurately perceive when my heart is beating fast.” The scale includes of 21 items rated on a scale from strongly agree (5) to strongly disagree (1), with total scores ranging from 21 to 105. Higher scores show greater self-reported interoceptive accuracy. A Dutch version of the IAS (IAS-D), that has good internal consistency (α = 0.89), was used (Appendix A; Mulder et al., In preparation). English and German versions of the IAS have been shown to possess good to excellent internal consistency (α = 0.84 to 0.90, ω = 0.81 to 0.88; Brand et al., 2023; Gabriele et al., 2022; Murphy et al., 2020; Tünte et al., 2024).

#### 2.2.2. Self-reported interoceptive attention

The Interoceptive Attention Scale (IATS; Gabriele et al., 2022) is a questionnaire constructed to quantify the extent to which internal signals are the object of one’s attention. The signals exactly match the signals included in the IAS. An example of a question in this questionnaire is “Most of the time my attention is focused on whether my heart is beating fast.” The IATS comprises 21 items rated from strongly agree (5) to strongly disagree (1), with total scores ranging from 21 to 105. Higher scores indicate greater self-reported attention to internal signals. A Dutch version of the IATS (IATS-D), with excellent internal consistency (α = 0.94), was used (Appendix B; Mulder et al., In preparation). The internal consistency of the IATS has been shown to be good to excellent in English and German speaking samples (α = 0.91, ω = 0.85 to 0.92; Gabriele et al., 2022; Tünte et al., 2024).

#### 2.2.3. Physical activity

The International Physical Activity Questionnaire (IPAQ; Booth, 2000) covers four domains of physical activity: work-related, transportation, housework/gardening, and leisure-time activity. Additionally, the questionnaire includes questions about time spent sitting as an indicator of sedentary behavior. In this study the fourth domain (“Physical activities related to sports and leisure”) was used to determine the time spent on physical activity. Number of days per week and time spent per day in walking, moderate, and vigorous intensity physical activities were recorded. Hours per week reported in walking-, moderate-, and vigorous-intensity activities were used as outcome measures. Additionally, participants were asked which physical activities or sport(s) they generally participate in, as well as years of experience in these activities. Physical activities and sports were categorized in team (i.e., open-skilled activities were movement, task, and environmental requirements are dynamically changing) or individual (i.e., closed-skill activities were movement forms are fixed, and environmental and task constraints are constant) activities (Heilmann et al., 2022). Participants were categorized as inactive when their weekly physical activity did not exceed 150 minutes, as per the guidelines of the Dutch Ministry of Health, Welfare and Sport, and the Health Council of the Netherlands (Health Council of the Netherlands, 2017).

#### 2.2.4. Self-regulation

The Short Self-Regulation Questionnaire (SSRQ; Carey et al., 2004) is a 31-item measure of the ability to regulate behavior in order to achieve desired future outcomes. The SSRQ is a short form of the original Self-Regulation Questionnaire. Response options range from strongly disagree (1) to strongly agree (5), with total scores ranging from 31 to 155. Example of questions in this questionnaire are “I usually keep track of my progress towards my goals” and “I have trouble making up my mind about things.” Higher scores indicate better self-regulatory skills. The SSRQ was not available in Dutch, therefore the questionnaire was translated by an organization specialized in medical translations (SSRQ-D; Appendix C). Further changes were made by conducting think-aloud interviews, following guidelines for cross-cultural health research (Sousa & Rojjanasrirat, 2011). The English SSRQ has an alpha of 0.92, and correlates highly to the original Self-Regulation Questionnaire (r = 0.96; Carey et al., 2004).

### 2.3. Statistical analyses and software

All analyses were performed using a custom script written in Python (version 3.12), using the scipy (version 1.14) and statsmodels (version 0.14) packages. Normality of data for all measures was checked beforehand, by visually examining the distribution of data using histograms and Q-Q plots, and performing the Shapiro-Wilk test. Missing values were managed using the Multiple Imputation by Chained Equations with random forests (MICEforest version 6.0) package in Python, generating ten imputed datasets and running 20 iterations.

The sample consisted mainly of middle aged adults, who participated less in vigorous intensity and team sports physical activities. Therefore, three types of regression analyses were considered and compared based on model fit statistics, to counter these skewed group sizes: multivariate linear regression analyses with and without log-normalization of dependent and independent variables, and Bayesian regression analyses. Based on model fit statistics, multivariate regression analyses with log-normalization were adopted, with the IAS-D and IATS-D as dependent variables, and the three IPAQ categories (walking, moderate, and vigorous intensity activities), years of experience in the physical activity generally participated in, and SSRQ-D as independent variables. Additionally, interaction effects between self-regulation and walking-, moderate-, and vigorous intensity activities were explored. Age, sex, and socio-economic status (SES) were entered into the final regression model to check for potential confounding effects, using a stepwise method and keeping those confounders that improved the model. Additionally, we explored sensitivity analyses with groups split on the median of the self-regulation scores, the results of which are shown in Appendix D. Coefficients were interpreted using Cohen’s f^2^: coefficients of 0.02 were considered small, 0.15 medium, and 0.35 large (Cohen, 2013). Alpha was set at 0.05 a priori. Adjusted significance based on Bonferroni corrections was set at 0.0071 a priori.

## 3. Results

### 3.1. Descriptive statistics

Responses from 1242 participants were collected. After removing outliers (values outside 3SD for the interoception, hours of physical activity, and years of physical activity experience variables), 1178 participants remained. The average age of the participants was 52.13 ± 16.78, and 52% were female. In the current sample, about 28% of the participants did not meet Dutch physical activity guidelines, which is lower than the national average of 56%. Furthermore, only a small percentage (about 6%) of the participants indicated generally participating in team sport activities. Additional descriptives of the participants are presented in Table 1.

**Table 1.** Descriptive statistics for continuous variables in mean ± SD and categorical variables in count (%), per physical activity type (inactive, participating in team sports, or participating in individual sports).

|  | Total<br>(n=1178) | Inactive<br>(n=325) | Team<br>(n=72) | Individual<br>(n=781) | p* |
| --- | --- | --- | --- | --- | --- |
| *Continuous variables* |  |  |  |  |  |
| Age | 52.13 ± 16.78 | 53.87 ± 17.14 | 46.94 ± 18.33 | 51.89 ± 16.39 | 0.005<sup>a,c</sup> |
| Dutch Interoceptive Accuracy Scale total score | 81.66 ± 9.62 | 80.91 ± 9.84 | 84.58 ± 10.49 | 81.70 ± 9.40 | 0.013<sup>a,c</sup> |
| Dutch Interoceptive Attention Scale total score | 57.63 ± 16.63 | 58.17 ± 15.86 | 61.90 ± 20.22 | 57.01 ± 16.54 | 0.045 |
| Walking activities (hours per week) | 3.75 ± 4.17 | 0.66 ± 0.72 | 3.56 ± 2.37 | 5.06 ± 4.46 | <0.001<sup>a,b,c</sup> |
| Moderate intensity activities (hours per week) | 2.01 ± 3.29 | 0.17 ± 0.43 | 1.17 ± 2.66 | 2.85 ± 3.66 | <0.001<sup>a,b,c</sup> |
| Vigorous intensity activities (hours per week) | 1.33 ± 2.52 | 0.08 ± 0.29 | 1.44 ± 2.41 | 1.84 ± 2.85 | <0.001<sup>a,b</sup> |
| Total active hours per week | 7.10 ± 6.92 | 0.92 ± 0.85 | 6.17 ± 4.60 | 9.75 ± 6.86 | <0.001<sup>a,b,c</sup> |
| Physical activity experience (years) | 13.39 ± 15.39 | 4.31 ± 11.58 | 16.73 ± 12.75 | 16.86 ± 15.47 | <0.001<sup>a,b</sup> |
| Dutch Short Self-Regulation Questionnaire total score | 109.36 ± 13.50 | 106.02 ± 13.85 | 110.96 ± 12.81 | 110.61 ± 13.19 | <0.001<sup>a,b</sup> |
| *Categorical variables* |  |  |  |  |  |
| Sex |  |  |  |  | 0.008<sup>c</sup> |
| Female | 612 (52%) | 183 (56%) | 26 (36%) | 403 (52%) |  |
| Male | 566 (48%) | 142 (44%) | 46 (64%) | 378 (48%) |  |
| SES |  |  |  |  | <0.001<sup>c</sup> |
| Low | 539 (46%) | 183 (56%) | 26 (36%) | 330 (42%) |  |
| High | 639 (54%) | 142 (44%) | 46 (64%) | 451 (58%) |  |

\*P-values based on ANOVA for continuous variables and Chi-Square tests for categorical variables, with post-hoc analyses. Significant difference between: <sup>a</sup>Inactive vs. Team; <sup>b</sup>Inactive vs. Individual; <sup>c</sup>Team vs. Individual.

### 3.2. Hypotheses testing

More walking hours per week (β = 0.009, SE = 0.004, p = 0.028) was weakly related to better interoceptive accuracy in the univariate analyses (Table 2), but moderate- and vigorous-intensity physical activity was not. Next, self-regulation was found to be significant in the univariate model (β = 0.282, SE = 0.026, p < 0.001), model 1 (β = 0.284, SE = 0.026, p < 0.001), and the final model (β = 0.262, SE = 0.027, p < 0.001), with medium to large effect sizes. It was found that with an increase in self-regulation scores, interoceptive accuracy increased as well. The confounders sex (β = −0.034, SE = 0.017, p = 0.043), age (β = 0.024, SE = 0.010, p = 0.015), and SES (β = 0.072, SE = 0.017, p < 0.001) were found to be significant determinants in the univariate model. The addition of these confounders improved the fit of the final model, refining the coefficients of the independent variables but not changing their directions. No significant interaction terms were found.

**Table 2.** Regression analyses with log-normalization for the Dutch Interoceptive Accuracy Scale. Model 1 includes the independent variables; the final model includes the independent, and confounding variables that significantly improved the model. Significant findings are marked with *, significant findings after Bonferroni corrections are marked with †.

| Variable | Univariate Beta<br>(95% CI) | Model 1 Beta<br>(95% CI) | Final Model Beta<br>(95% CI) |
| --- | --- | --- | --- |
| Walking<br>(Hours per week) | 0.009*<br>(0.001 - 0.017) | 0.004<br>(-0.004 - 0.013) | 0.005<br>(-0.004 - 0.014) |
| Moderate intensity activities<br>(Hours per week) | 0.005<br>(-0.004 - 0.013) | 0.002<br>(-0.007 - 0.011) | 0.001<br>(-0.008 - 0.010) |
| Vigorous intensity activities<br>(Hours per week) | 0.002<br>(-0.007 - 0.011) | -0.006<br>(-0.015 - 0.003) | -0.006<br>(-0.016 - 0.003) |
| Physical activity experience<br>(years) | 0.003<br>(-0.002 - 0.008) | -0.002<br>(-0.008 - 0.004) | -0.003<br>(-0.009 - 0.003) |
| Self-regulation | 0.282*†<br>(0.232 - 0.332) | 0.284*†<br>(0.233 - 0.335) | 0.262*†<br>(0.209 - 0.314) |
| Sex | -0.034*<br>(-0.067 - -0.001) |  | -0.010<br>(-0.042 - 0.023) |
| Age | 0.024*<br>(0.005 - 0.043) |  | 0.026*<br>(0.005 - 0.046) |
| SES | 0.072*†<br>(0.039 - 0.105) |  | 0.050*†<br>(0.015 - 0.084) |

Physical activity experience was a significant determinant of interoceptive attention (Table 3) in the univariate model (β = −0.017, SE = 0.007, p = 0.013), model 1 (β = −0.019, SE = 0.008, p = 0.017), and the final model (β = −0.022, SE = 0.008, p = 0.005), with small effect sizes. Additionally, it was found that self-regulation was a significant determinant of interoceptive attention in the univariate model (β = −0.147, SE = 0.070, p = 0.036), showing a medium effect size. Sex (β = −0.177, SE = 0.044, p < 0.001), age (β = 0.095, SE = 0.025, p < 0.001), and SES (β = −0.321, SE = 0.043, p < 0.001) were significant in the univariate model, and added to the final model as they improved the fit of the model. Their addition did not change the direction of the independent variables. No significant interaction terms were found, but the self-regulation and vigorous intensity activity interaction approached significance (p = 0.059).

**Table 3.** Regression analyses with log-normalization for the Dutch Interoceptive Attention Scale. Model 1 includes the independent variables; the final model includes the independent, and confounding variables that significantly improved the model. Significant findings are marked with *, significant findings after Bonferroni corrections are marked with †.

| Variable | Univariate Beta<br>(95% CI) | Model 1 Beta<br>(95% CI) | Final Model Beta<br>(95% CI) |
| --- | --- | --- | --- |
| Walking<br>(Hours per week) | 0.001<br>(-0.021 - 0.022) | 0.016<br>(-0.008 - 0.040) | 0.021<br>(-0.002 - 0.043) |
| Moderate intensity activities<br>(Hours per week) | -0.012<br>(-0.034 - 0.010) | -0.001<br>(-0.026 - 0.023) | -0.002<br>(-0.025 - 0.022) |
| Vigorous intensity activities<br>(Hours per week) | -0.003<br>(-0.027 - 0.021) | 0.005<br>(-0.020 - 0.030) | 0.014<br>(-0.011 - 0.039) |
| Physical activity experience<br>(years) | -0.017*<br>(-0.030 - -0.003) | -0.019*<br>(-0.035 - -0.003) | -0.022*†<br>(-0.037 - 0.007) |
| Self-regulation | -0.147*<br>(-0.285 - -0.010) | -0.138<br>(-0.279 - 0.002) | -0.066<br>(-0.206 - 0.075) |
| Sex | -0.177*†<br>(-0.263 - -0.091) |  | -0.186*†<br>(-0.274 - -0.099) |
| Age | 0.095*†<br>(0.046 - 0.145) |  | 0.034<br>(0.020 - 0.088) |
| SES | -0.321*†<br>(-0.406 - -0.236) |  | -0.308*†<br>(-0.400 - -0.215) |

## 4. Discussion

The current study examined how varying intensities of physical activity (e.g., walking, moderate intensity, and vigorous intensity physical activity) are associated with self-reported interoceptive accuracy and attention. Additionally, this study investigated the influence of individual (i.e., years of physical activity experience and self-regulatory skills) factors on self-reported interoceptive accuracy and attention. Results showed that more walking hours per week (however, only in the univariate model) and higher levels of self-regulation scores were related to higher self-reported interoceptive accuracy, while more physical activity experience and higher levels of self-regulation were related to lower levels of interoceptive attention. Moderate and vigorous physical activity hours per week were not associated with self-reported interoceptive accuracy and attention.

The findings that walking hours per week was related to better interoceptive accuracy, but moderate and vigorous intensity physical activity hours per week were not, and that years of physical activity experience was related to interoceptive attention, might be explained by adaptation or habituation effects related to adaptive (i.e., balanced bodily awareness beneficial for health-promoting behavior) forms of interoception (Aaron et al., 2020; Trevisan et al., 2023). Becoming more accustomed to bodily changes induced by physical activity reduces the perceived sensitivity to arousal signals, which allows more attentional resources to be put towards external stimuli (Marshall et al., 2018; Wallman-Jones et al., 2023). The same adaptations may occur over longer periods of time, resulting in interoceptive attention decreasing with more years of physical activity experience. Or in other words, with more experience in perceiving internal signals during physical activity, individuals may start to trust their body more and shift their attentional focus from internal to external. These habituation effects relate to the central command network and ratings of perceived exertion: during physical activity, signals from the brain, accompanied by memories of prior experiences, help to manage energy expenditure and the intensity of physical exertion through interoceptive predictions. Ratings of perceived exertion are generated by interoceptive processes that compare the expected sensations with incoming bodily signals (Brevers et al., 2024; McMorris et al., 2018). During lower intensity physical activities, these cognitive control strategies are easier to maintain, future interoceptive states are easier to predict, and there are fewer interoceptive prediction errors. This allows for better interoceptive regulation and the adaptation to physical activity intensities so homeostasis is not threatened (McMorris et al., 2018; Seabury et al., 2023). There may be a bidirectional relationship between interoception and physical activity, where increased exposure to physical activity enhances the accuracy of internal models, which, in turn, helps better manage exertion (Seabury et al., 2023). In turn, increased accuracy may result in decreased attention to bodily signals because of an increase in bodily trust. This could explain the findings on hours spent on walking activities and years of physical activity experience.

The finding that hours per week engaged in vigorous intensity physical activity did not significantly determine self-reported interoceptive accuracy and attention scores is might be the result of higher internal noise during these types of physical activity (Koteles et al., 2020; Wallman-Jones et al., 2022). In contrast to lower intensity activities, during vigorous intensity physical activity it becomes more difficult to make top-down interoceptive predictions: cognitive control strategies become less effective, future states are harder to predict, and interoceptive prediction errors become more prevalent (Seabury et al., 2023). When physical activity intensifies, interoceptive cues call for more attention, decreasing the availability of attentional resources (Razon et al., 2014; Wallman-Jones et al., 2022). Instead, attention may shift towards associative imagery (e.g., increasing engagement with physical movement and bodily experiences, and enhancing motivation to cope with effort perceptions) in order to adapt to physiological sensations and extend task performance and duration (Razon et al., 2010). Additionally, intense physical activity can negatively impact emotional well-being due to the body’s physical state (Ekkekakis, 2003). These emotions (i.e., affective responses) combined with other internal physiological noise, compete with interoceptive signals, and make it harder to distinguish these feelings and sensations from one another. This may explain why in the current study hours per week participated in vigorous intensity physical activity did not determine scores on the IAS-D and IATS-D.

In line with previous studies (Füstös et al., 2013; Georgiou et al., 2015; Herbert et al., 2007; Marshall et al., 2018), the current study found a positive significant association between self-regulatory skills and interoceptive accuracy. In addition to these earlier findings the current study showed a significant negative association between self-regulation and interoceptive attention. It may be that individuals with higher self-regulation scores do not need to pay as much attention to accurately perceive internal signals, meaning they are more efficient. Affective responses to physical activity come from the combination of cognitions originating in the frontal cortex, such as self-regulation, and interoceptive signals about the physiological state of the body (Ekkekakis, 2009). Increased interoceptive accuracy improves self-regulatory skills, which in turn allows better management of attention and emotions (Seabury et al., 2023). For example, individuals who are good at perceiving and interpreting bodily signals may also be good at self-regulation. Or vice versa: if bodily signals are not interpreted correctly, it becomes more difficult to self-regulate. It could be that with better interoceptive accuracy and self-regulation comes more trust in one’s bodily signals, resulting in less attention paid to those sensations.

The novelty of the current study is that it investigated time spent in multiple intensities (i.e., walking, moderate, and vigorous) of physical activity in two separate interoceptive domains (i.e., accuracy and attention), providing more information by tapping into different aspects of interoception (Schoeller et al., 2025). To the best of our knowledge, previous research has mainly focused on the effect of acute exercise protocols on interoception in a lab-setting, as well as in single interoceptive domains. Thus, the current study adds to existing information by investigating the association between the intensity and amount of physical activity individuals participate in during their leisure time and several interoceptive domains, which might result in a better comprehension of how interoception is used in daily physical activities.

There were some limitations to this study. First, we should note that conclusions on the relation between hours per week spent on different intensities physical activity and self-reported interoception should be interpreted with caution. The questionnaires we used are of a more general nature and were not directly linked to different physical activity intensities (i.e., we did not ask participants to take specific physical activities or intensities in mind when filling out the questionnaires). Therefore, our conclusions may indicate possible mechanisms, but we are not able to mention causal relations. However, the outcomes of this explorative study may provide a starting point for future research on the associations between physical activity intensities, self-regulation, and interoceptive accuracy and attention. Second, participants aged 18 years and older from the general Dutch population were included in the study, which led to the participants ranging in age from 18 to 96 with a mean of 52. This on average middle-aged sample has a different physical activity profile compared to for example young adults, which means they took part less in vigorous intensity activities than their younger counterparts. Although we attempted to address this using log-normalization, the results of this study may be limited and not fully generalizable across age groups. Finally, the current study relied solely on self-report measures. There are known limitations to self-report measures, such as cultural differences, recall bias, and misinterpretation of questions (Corder & van Sluijs, 2010). For the IPAQ specifically, there is also the tendency to overestimate actual physical activity levels (Steene-Johannessen et al., 2016). Additionally, because of the complexity of interoception, relationships between self-report and objective interoceptive measures must also be considered in order to paint a complete picture of the association between physical activity and interoception (Schoeller et al., 2025). However, the current results do provide insight into the association between interoception and physical activity, and can be used as a starting point for further experimental research.

More research is needed to better understand the complex relations between physical activity, self-regulation, and interoception. Based on the results of the current study, this research should focus on possible mechanisms underlying these relations. An interesting next step for research would be to investigate whether individuals with better self-regulatory skills are more physically active because of these skills, whether this leads to them being able to more accurately perceive their internal bodily signals, and subsequently pay less attention to internal bodily signals because of improved bodily trust. Insights into these relations and mechanisms are extremely relevant for promoting health behavior, as it may help individuals filter the most relevant interoceptive information and engage in more adaptive forms of interoception (i.e., balanced bodily awareness beneficial for health-promoting behavior). Additionally, future research should pay more attention to the association between different physical activity types (e.g., cardio versus strength, team versus individual, or organized versus non-organized) and different interoceptive domains and modalities, considering how different populations associate with specific interoceptive profiles (Schoeller et al., 2025). Insights into these differences could allow for optimization of specific physical activity and interoception interventions for specific target populations. Finally, to understand how knowledge on the relation between different physical activity intensities and interoceptive domains can be used to inform lifestyle interventions, more longitudinal research is needed. Longitudinal research may allow us to investigate the timing of physical activity and interoceptive interventions (i.e., how long it takes for these interventions to show effects) as well as examine the bidirectional relationship between physical activity and interoception.

## 5. Conclusion

Walking hours per week associated with an increase in interoceptive accuracy, while more years of PA experience decreased interoceptive attention. At lower intensity physical activity, adaptation or habituation effects reduce perceived sensitivity to internal signals, meaning attention is shifted more towards external stimuli. Similar adaptations may occur over longer periods of time, resulting in interoceptive attention decreasing with more years of physical activity experience. Better self-regulatory skills were associated with increased interoceptive accuracy, and decreased interoceptive attention. It could be that with better interoceptive accuracy and self-regulation comes more trust in one’s bodily signals, resulting in less attention paid to those sensations. Being able to filter relevant interoceptive information may be extremely important in health behavior. An interesting next step for researchers is to investigate whether individuals with better self-regulation are therefore more physically active, resulting in increased interoceptive accuracy and decreased interoceptive attention.

## Data Availability

All data produced in the present study are available upon reasonable request to the authors.

## 6. Other information

### Author contributions

JM, JKJ, and MEG conceptualized and designed the study. JM performed data collection. JM and JKJ conducted statistical analyses. JM, JKJ, JdV, and MEG contributed to the interpretation of the results. JM drafted the initial manuscript, with JKJ, JdV, and MEG providing critical revisions and refining the final manuscript. All authors reviewed and approved of the final version of the manuscript.

### Data availability

Questionnaires in Dutch, data, and analysis scripts will be made available by the corresponding author upon reasonable request.

### Declaration of generative AI and AI-assisted technologies in the writing process

During the preparation of this work the author(s) used ChatGPT in order to help with writing the code for the statistical analyses. After using this tool/service, the author(s) reviewed and edited the content as needed and take(s) full responsibility for the content of the published article.

## 8. Appendix

**Appendix A: Dutch IAS (IAS-D)**

Made available by contacting the corresponding author upon reasonable request.

**Appendix B: Dutch IATS (IATS-D)**

Made available by contacting the corresponding author upon reasonable request.

**Appendix C: Dutch SSRQ (SSRQ-D)**

Made available by contacting the corresponding author upon reasonable request.

