## Supplemental File 1 for "Associations Between Physical Activity Intensity and Experience, Self-Regulation, and Self-Reported Interoceptive Accuracy and Attention"

### Supplemental file 1: Sensitivity analyses with groups split by SSRQ-D median

*Supplemental Table 1: Regression analyses with log-normalization for the Dutch Interoceptive Accuracy Scale. Model 1 includes the independent variables; the final model includes the independent, and confounding variables that significantly improved the model. Significant findings are marked with *, significant findings after Bonferroni corrections are marked with †.*

| **Variable** | **Univariate Beta (95% CI)** | | **Model 1 Beta (95% CI)** | | **Final model Beta (95% CI)** | |
|---|---|---|---|---|---|---|
| | *Low SSRQ-D Group* | *High SSRQ-D Group* | *Low SSRQ-D Group* | *High SSRQ-D Group* | *Low SSRQ-D Group* | *High SSRQ-D Group* |
| Walking | 0.008 | 0.001 | 0.008 | 0.002 | 0.008 | 0.002 |
| (Hours per week) | (-0.003 – 0.020) | (-0.011 – 0.012) | (-0.006 – 0.019) | (-0.010 – 0.013) | (-0.004 – 0.020) | (-0.009 – 0.014) |
| Moderate intensity activities | 0.004 | 0 | 0.004 | 0.001 | 0 | 0.001 |
| (Hours per week) | (-0.008 – 0.015) | (-0.011 – 0.011) | (-0.012 – 0.014) | (-0.011 – 0.013) | (-0.013 – 0.013) | (-0.011 – 0.012) |
| Vigorous intensity activities | -0.002 | -0.004 | -0.002 | -0.004 | -0.005 | -0.008 |
| (Hours per week) | (-0.016 – 0.012) | (-0.016 – 0.008) | (-0.021 – 0.008) | (-0.016 – 0.008) | (-0.020 – 0.009) | (-0.020 – 0.004) |
| Physical activity experience | 0.004 | -0.004 | 0.004 | -0.005 | 0 | -0.005 |
| (years) | (-0.003 – 0.010) | (-0.011 – 0.004) | (-0.007 – 0.009) | (-0.013 – 0.003) | (-0.008 – 0.008) | (-0.013 – 0.003) |
| Self-regulation | 0.142 | 0.677 | 0.142 | 0.680 | 0.125 | 0.641 |
| | (0.051 – 0.233)*† | (0.530 – 0.824)*† | (0.048 – 0.232)*† | (0.532 – 0.827)*† | (0.033 – 0.218)* | (0.492 – 0.791)*† |
| Sex | -0.006 | -0.048 | | | 0.011 | -0.037 |
| | (-0.051 – 0.040) | (-0.094 - -0.003)* | | | (-0.035 – 0.057) | (-0.081 – 0.006) |
| Age | 0.032 | 0.007 | | | 0.038 | 0.046 |
| | (0.006 – 0.058)* | (-0.019 – 0.034) | | | (0.010 – 0.067)* | (0 – 0.093)* |
| SES | 0.024 | 0.076 | | | 0.035 | 1.362 |
| | (-0.022 – 0.069) | (0.029 – 0.123)*† | | | (-0.013 – 0.083) | (0.646 – 2.078)*† |

*Supplemental Table 2: Regression analyses with log-normalization for the Dutch Interoceptive Attention Scale. Model 1 includes the independent variables; the final model includes the independent, and confounding variables that significantly improved the model. Significant findings are marked with *, significant findings after Bonferroni corrections are marked with †.*

| **Variable** | **Univariate Beta (95% CI)** | | **Model 1 Beta (95% CI)** | | **Final model Beta (95% CI)** | |
|---|---|---|---|---|---|---|
| | *Low SSRQ-D Group* | *High SSRQ-D Group* | *Low SSRQ-D Group* | *High SSRQ-D Group* | *Low SSRQ-D Group* | *High SSRQ-D Group* |
| Walking | 0.003 | 0.005 | 0.014 | 0.018 | 0.019 | 0.020 |
| (Hours per week) | (-0.025 – 0.032) | (-0.028 – 0.038) | (-0.017 – 0.046) | (-0.018 – 0.053) | (-0.012 – 0.050) | (-0.014 – 0.054) |
| Moderate intensity activities | -0.014 | -0.006 | -0.007 | 0.006 | -0.008 | 0.006 |
| (Hours per week) | (-0.044 – 0.016) | (-0.037 – 0.026) | (-0.041 – 0.027) | (-0.030 – 0.041) | (-0.041 – 0.025) | (-0.028 – 0.040) |
| Vigorous intensity activities | 0.012 | -0.008 | 0.018 | -0.005 | 0.019 | 0.015 |
| (Hours per week) | (-0.023 – 0.047) | (-0.042 – 0.026) | (-0.019 – 0.054) | (-0.040 – 0.031) | (-0.017 – 0.055) | (-0.021 – 0.050) |
| Physical activity experience | -0.013 | -0.017 | -0.017 | -0.022 | -0.016 | -0.031 |
| (years) | (-0.030 – 0.004) | (-0.037 – 0.004) | (-0.037 – 0.004) | (-0.045 – 0.002) | (-0.036 – 0.005) | (-0.054 – -0.008)* |

| | | | | | | |
|---|---|---|---|---|---|---|
| Self-regulation | -0.030<br>(-0.263 – 0.202) | -0.119<br>(-0.564 – 0.327) | -0.030<br>(-0.265 – 0.204) | -0.155<br>(-0.561 – 0.332) | 0.005<br>(-0.228 – 0.237) | 0.039<br>(-0.398 – 0.476) |
| Sex | -0.150<br>(-0.265 – -0.036)* | -0.217<br>(-0.346 - -0.087)*† | | | -0.161<br>(-0.275 – -0.048)*† | -0.195<br>(-0.328 – -0.063)*† |
| Age | 0.037<br>(-0.030 – 0.104) | 0.160<br>(0.086 – 0.235)*† | | | -0.252<br>(-0.366 – -0.137)*† | 0.089<br>(0.007 – 0.170)* |
| SES | -0.238<br>(-0.352 – -0.125)*† | -0.394<br>(-0.526 – -0.262)*† | | | 4.399<br>(3.328 – 5.470)*† | -0.361<br>(-0.504 – -0.219)*† |
